# Machine learning identifies a COVID-19-specific phenotype in university students using a mental health app

**DOI:** 10.1101/2022.12.07.22283234

**Authors:** Artur Shvetcov, Alexis Whitton, Suranga Kasturi, Wu-Yi Zheng, Joanne Beames, Omar Ibrahim, Jin Han, Leonard Hoon, Kon Mouzakis, Sunil Gupta, Svetha Venkatesh, Helen Christensen, Jill Newby

**Affiliations:** Black Dog Institute, UNSW Sydney, NSW, Australia; Applied Artificial Intelligence Institute, Deakin University, Geelong, VIC, Australia

## Abstract

Advances in smartphone technology have allowed people to access mental healthcare via digital apps from wherever and whenever they choose. University students experience a high burden of mental health concerns. Although these apps improve mental health symptoms, user engagement has remained low. Studies have shown that users can be subgrouped based on unique characteristics that just-in-time adaptive interventions (JITAIs) can use to improve engagement. To date, however, no studies have examined the effect of the COVID-19 pandemic on these subgroups. Here, we use machine learning to examine user subgroup characteristics across three COVID-19-specific timepoints: during lockdown, immediately following lockdown, and three months after lockdown ended. We demonstrate that there are three unique subgroups of university students who access mental health apps. Two of these, with either higher or lower mental well-being, were defined by characteristics that were stable across COVID-19 timepoints. The third, situational well-being, had characteristics that were timepoint-dependent, suggesting that they are highly influenced by traumatic stressors and stressful situations. This subgroup also showed feelings and behaviours consistent with burnout. Overall, our findings clearly suggest that user subgroups are unique: they have different characteristics and therefore likely have different mental healthcare goals. Our findings also highlight the importance of including questions and additional interventions targeting traumatic stress(ors), reason(s) for use, and burnout in JITAI-style mental health apps to improve engagement.

## Introduction

Over the past decade, advances in digital technology have driven rapid changes in the way mental health care is accessed and delivered. In particular, improvements in smartphone technology have afforded opportunities to design mental health applications (‘apps’) that can be accessed wherever and whenever a user chooses, and that can deliver strategies *in situ* to assist users in coping with stressors as they occur in real time. As they are easily and rapidly accessible by anyone with a smartphone, smartphone-based mental health apps overcome many of the barriers individuals encounter when trying to access face-to-face mental health services, including long wait times for appointments, high cost, lack of time, and concerns about privacy ^1^. As a result, smartphone-based mental health interventions have grown in popularity in recent years, yet our understanding of who uses these interventions, and why, remains limited.

Given the ubiquity of smartphones among young adults, smartphone-based mental health apps have received considerable interest as a means for addressing the mental health needs of university student populations. University students experience a disproportionate burden of common mental health symptoms, such as anxiety and depression, compared to both age-matched peers and to the general population ^2, 3, 4, 5^. Recent studies indicate that the number of university students experiencing common mental health conditions rose substantially during the COVID-19 pandemic ^6, 7, 8, 9, 10^. Although most universities provide on-campus counselling and other mental health services, the demand for these services far exceeds service availability, meaning that most university students with mental health conditions go untreated.

Several smartphone-based apps have been shown to improve symptoms of depression and anxiety and increase well-being in university students ^11, 12^. However, a common shortcoming of these apps is that user engagement is often low ^1, 13, 14, 15^. Although tailoring app content can improve app engagement, even tailored apps have been found to have substantial rates of attrition ^15^, suggesting that other factors may need to be considered in order to enhance app engagement.

University students access mental health apps for a range of different reasons ^16^ and one relatively underexplored factor that may determine levels of app engagement is an individual’s motivation for app use. Although the body of evidence is still limited, research characterising the distinctive features of subgroups of mental health app users indicates that there are typically four subgroups of users that can be differentiated based on sex, likelihood of having mental health problems and accessing mental health services, engagement in health-related behaviour (e.g. fruit consumption), and type of mental health concern (e.g. depression vs. distress) ^17, 18^. Prior studies have shown that information about these user subgroups can provide a valuable means for enhancing engagement with mental health apps. For example, in a sample of university students who took part in online mental health screening, subgroup-tailored mental health feedback – personalized using cluster analysis on multidimensional aspects of mental health – was found to increase university students’ engagement with their feedback and boost their mental health literacy ^19^.

Despite the clear benefits of subgrouping users, most studies to date have focused on subgrouping individuals using data on personal characteristics (e.g., age, gender, symptoms). However, situational factors, such as acute environmental stressors, can also have a powerful influence on an individual’s motivation to engage with mental health treatment ^20^, and further work is needed to better understand how user-specific situational factors can be used to tailor smartphone-based mental health interventions. The COVID-19 pandemic and its associated lockdowns have significantly affected the mental health of people around the world and particularly university students. University students reported to experience significant increases in depression, anxiety, suicidal thoughts, and perceived stress along with a worsened quality of life and mental well-being ^6, 7, 9, 10^. Further, COVID-19 lockdowns have been identified as a universal traumatic stressor ^21^. This presents a unique opportunity to examine both the stability of mental health app user subgroups and how a stressor influences these subgroups. More specifically, by comparing profiles of app users across the stages of COVID-19 lockdowns (i.e. during lockdown, immediately after lockdown, and well after lockdown) we can identify if major life stressors affect user subgroup characteristics. In doing so, we may be able to further improve the personalization of mental health apps thereby increasing user engagement.

In this study, we identified the effects of the COVID-19 pandemic and the associated stages of lockdown on the characteristics of university student mental health app user subgroups in Australia. We aimed to (1) determine and characterize different subgroups of university student mental health app users based on their responses to psychological surveys and (2) examine the influence that the COVID-19 pandemic had on the stability and characteristics of these user subgroups. To address this, we used a combination of unsupervised and supervised machine learning to identify user subgroup-defining characteristics across three COVID-19 pandemic timepoints: lockdown, immediately post-lockdown, and normal (at least 3 months past the lifting of lockdown restrictions).

## Method

### Study Design and Participants

The present analysis was performed using participant screening data collected in the context of an adaptive trial of a smartphone-based program (‘Vibe Up’) that delivers brief mental health interventions to Australian-based university students ^22^. Using an Artificial Intelligence (AI)-driven response adaptive randomization design, the trial used a series of sequential ‘mini trials’ to identify which of four, two-week smartphone interventions (mindfulness, physical activity, or sleep hygiene, with a mood monitoring intervention used as an active control condition) was the most effective for reducing symptoms of psychological distress in university students. All screening data was included irrespective of whether the participant went on to enroll in and/or complete the study. The study was approved by the University of New South Wales Human Research Ethics Committee, approval no. HC200466.

Participant screening data from the trial was divided into three separate datasets according to their COVID-19-related situational circumstances at the time of screening. The first dataset represented participants who were in COVID-19 lockdowns in Australian cities at the time of screening (lockdown timepoint). The second dataset consisted of participants who were screened immediately after the cessation of the COVID-19 lockdowns (post-lockdown timepoint). The third dataset represented those who were screened at least three months past the COVID-19 lockdowns (normal timepoint).

### Psychological Surveys Used to Identify User Characteristics

Prospective participants completed a battery of self-report questionnaires in the Vibe Up smartphone application as part of screening, and scores on these questionnaires were used as the features on which our machine learning models were trained. The questionnaires included the Abridged NIDA-Modified ASSIST Drug Screening Tool (AOD) ^23^, EuroQuol-5D 5-level version (EQ-5D-5L) ^24^, Kessler Psychological Distress Scale 10-item version (KTEN) ^25^, Use of Mental Health Care Services (MHS) ^26^, Perceived Stress Scale (PSS) ^27^, Physical and Mental Health (MED) ^22^, Suicidal Ideation Attributes Scale (SIDAS) ^28^, and Short Warwick Edinburgh Mental Well-being Scale (WBS) ^29^ (see Supplementary Methods for further details). Scores on all questionnaires were standardized by converting to z-scores prior to analyses. A series of demographic questions were also asked including self-identified gender, sex at birth, and sexual orientation.

### Analytic approach

A complete overview of the analytical approach adopted for the present study is shown in Figure 1.

**Figure 1.**
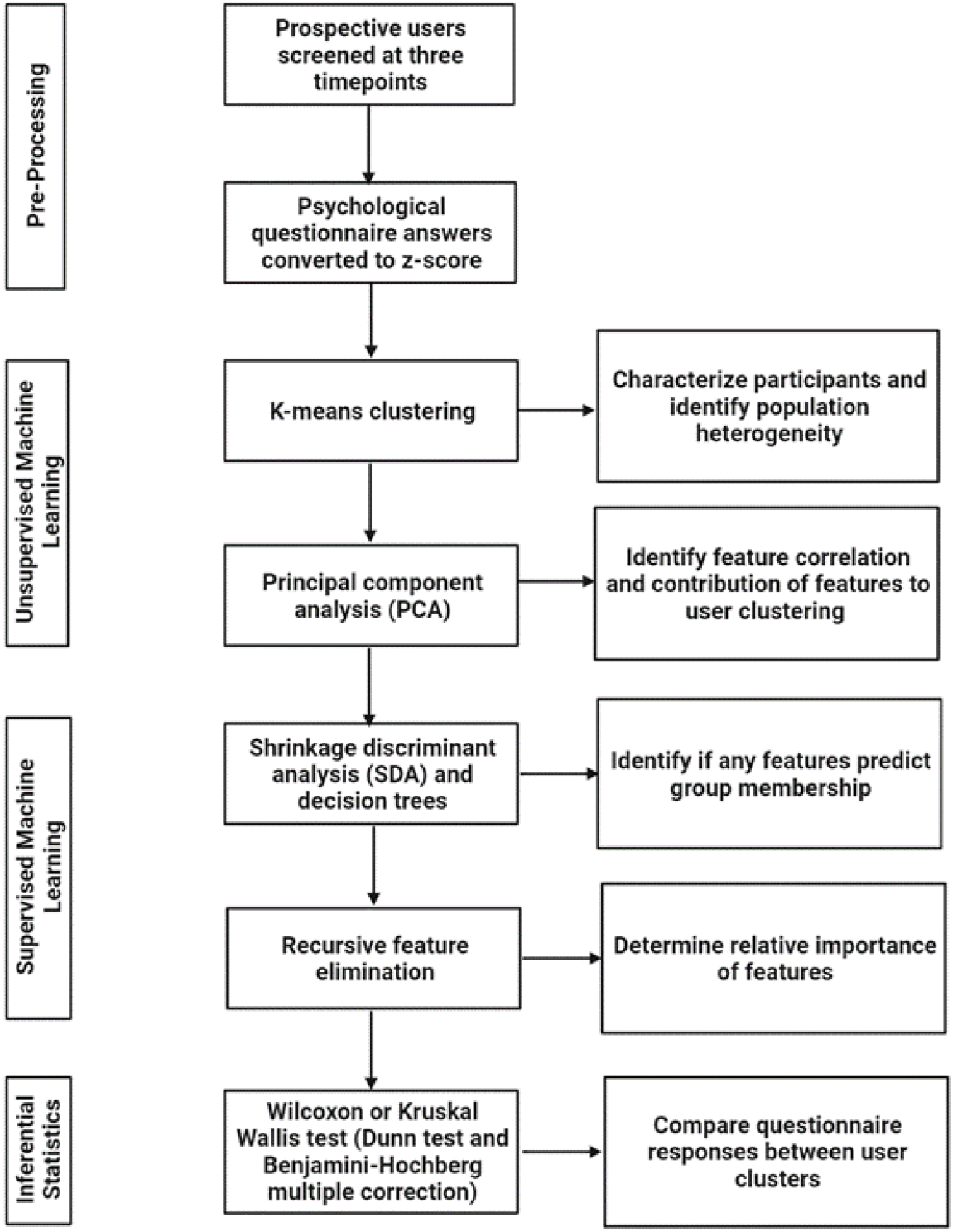
Summary of the method, statistical, and machine learning approaches used in the present study.

#### Unsupervised Machine Learning

A k-means clustering algorithm was used to characterize participants and to identify the degree of heterogeneity within the population. A principal component analysis (PCA) was then used to determine the potential correlations of the separate features (questionnaire responses) as well as the contribution of these features to the clustering of prospective participants. This was an important step to ensure that features that were strongly correlated with one another were not used in the supervised machine learning model as this affects the performance, even if the algorithm is robust ^30^. Clustering and PCA were performed in RStudio with R (3.6.3) using *base* and *factominer* / *factoextra* packages, respectively.

#### Supervised Machine Learning

To identify whether any of the features predicted cluster (subgroup) membership, we performed supervised machine learning using both shrinkage discriminant analysis and decision trees. We then performed model training/validation and testing on each of the three separate participant datasets (i.e., lockdown dataset, post-lockdown dataset, normal dataset), alternating which of the three datasets was used for training/validating or testing, such that all possible iterations were used (e.g., model 1: training = lockdown dataset, testing = post-lockdown dataset; model 2: training = post-lockdown dataset, testing = normal dataset, etc.). In every iteration, we used a 3-fold cross-validation repeated 5 times. In the event of an imbalance of classes in the output variable of the training dataset, upsampling was used where the less represented class was randomly sampled to make the number of samples equal to the more represented class. We used four metrics to assess the models’ performance: F1-score, recall, precision, and area under the curve (AUC). F1-score is the harmonic mean of the model’s precision and recall. Recall defines the model’s overall performance. Precision indicates how well the model discriminates the target of interest, in this case, the distinct cluster of prospective participants. The AUC indicates the model’s ability to distinguish between participant groups.

Following the use of our decision tree algorithm, we estimated the relative importance of features using recursive feature elimination (RFE). This algorithm starts with all features in a dataset and in every step removes one feature, refits the model, and recalculates the accuracy. As a result, each feature in a dataset can be ranked by its relative importance for model performance. Supervised machine learning was performed in RStudio with R (3.6.3) using the *caret* package.

#### Inferential Statistics

Inferential statistics were used to compare the questionnaire responses between the participant clusters. Wilcoxon tests were used for two-sample comparisons and Kruskal-Wallis tests followed by a post-hoc Dunn test were used for three sample comparisons. For all multiple tests, a Benjamini-Hochberg multiple correction was applied to adjust the *p* values and reduce the risk of a false positive. All inferential statistics were performed in RStudio with R (3.6.3) using the packages *base* and *fsa*.

#### Final Dataset Comparisons

Following all analyses, we compared the three datasets in order to identify the strength of the similarities and / or differences between them as well as how they changed over time. To do this, we first calculated the centers of the clusters for all three datasets and used a single graph to visualize the migration of the cluster centers to examine changes across the various stages of COVID-19 lockdown. We then calculated the Euclidean distances between datasets and built a hierarchical dendrogram. These analyses were performed in RStudio with R (3.6.3) using packages *base*, and *factominer / factoextra*.

## Results

### Dataset characteristics

Demographic characteristics of participants screened as part of the study are shown in Table 1. All participants screened were university students located in one of three Australian cities: Sydney, Melbourne, or Brisbane. The average age of participants screened was 22 (range 18 to 47), the majority identified their sex at birth as female, spoke English at home, and were domestic students. Approximately half had a previous mental health diagnosis. Between 20-30% of users had used online mental health services in the past 12 weeks (Table 1). There were no significant differences in the overall demographics recorded between the three COVID-19 timepoints.

**Table 1.** Demographics of participants screened

|  | **Lockdown**<br>(n = 308) | **Post-lockdown**<br>(n = 84) | **Normal**<br>(n = 81) |
| --- | --- | --- | --- |
| Age (average, ± SEM, and range) | 22.0 ± 3.2 (18-38) | 22.6 ± 4.2 (18-47) | 23.0 ± 4.0 (18-44) |
| Sex at birth | 91% female | 85% female | 91% female |
| Identify as LGBTQIA+ | 55% | 35% | 46% |
| Speak English at home | 94% | 96% | 88% |
| Domestic student | 93% | 96% | 93% |
| Previous mental health diagnosis | 55% | 53% | 55% |
| Used online mental health service in the past 12 weeks | 32% | 30% | 22% |

### General characterization of mental health app users shows three unique subgroups of users across lockdown, post-lockdown, and normal timepoints

To perform an initial characterization of the participants within the three timepoints, and the level of heterogeneity within the population, we employed unsupervised machine learning. Here, we performed a PCA for each timepoint, respectively. The PCAs demonstrated that each timepoint was characterized by three distinct clusters of participants (Figure 2).

**Figure 2.**
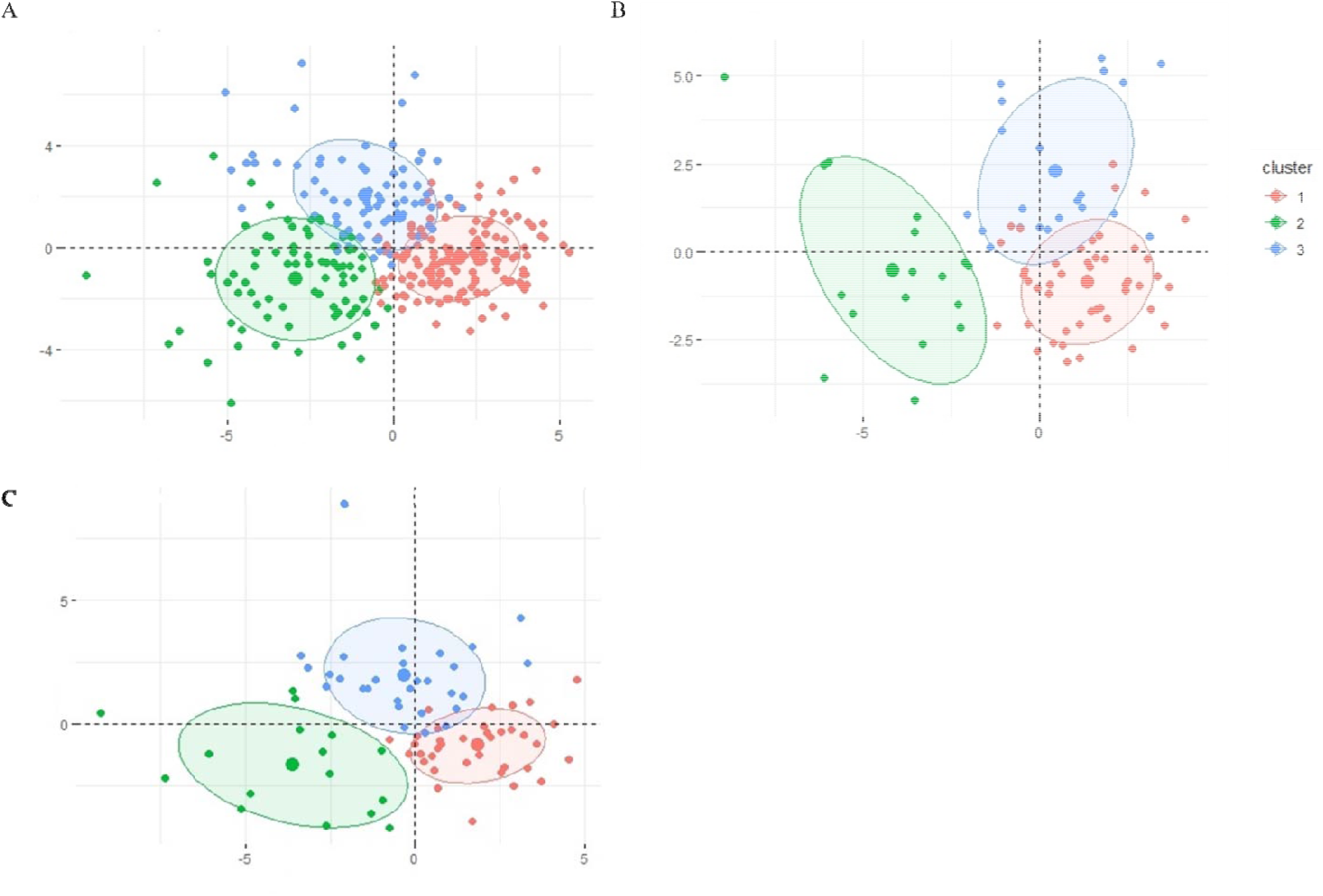
Principal component analysis (PCA) characterization of mental healthcare app users across three timepoints: (A) lockdown, (B) post-lockdown, and (C) normal.

Given that the PCA demonstrated the consistent presence of three clusters of app users across all timepoints, we then sought to establish if the features within each cluster were stable and could therefore be used to predict cluster membership. We used SDA for multiclass classification and alternated the training and testing datasets for each model. This demonstrated that cluster 1 was able to be predicted and had stable F1 and precision metrics irrespective of the timepoint used to train or test the model (performance metrics for each cluster are shown in Table 2). Cluster 2 was also able to be predicted across the various iterations of training and testing datasets. Unlike cluster 1, however, the model using post-lockdown as a training dataset and lockdown as a testing dataset performed poorly, showing an F1 of 0.62 and precision of only 0.53. This suggests that although cluster 2 is largely stable across the timepoints, there may be some subtle differences in defining features of this cluster between lockdown and post-lockdown. Compared to clusters 1 and 2, almost all the shrinkage discriminant analysis models had very poor performance, highlighting that cluster 3 was unable to be predicted. Here, only 2/6 models showed an adequate performance. Both models used combinations of lockdown and normal datasets for training and testing, respectively, suggesting that features defining cluster 3 may be similar across these two timepoints. The metrics of the remaining 4/6 showed that these models perform at, or below, chance levels.

**Table 2.**
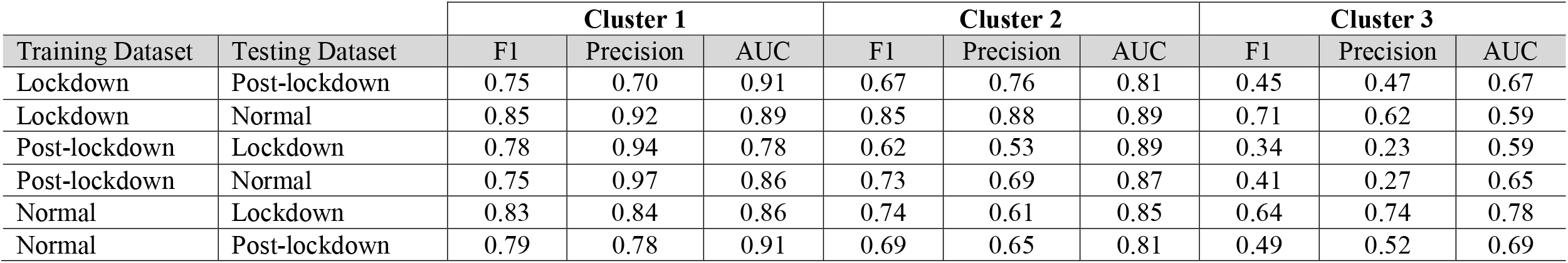
Performance metrics of the shrinkage discriminant analysis for multiclass classification of the three clusters of mental health app users. Metrics reported include F1, precision, and AUC.

| Training Dataset | Testing Dataset | Cluster 1 |  |  | Cluster 2 |  |  | Cluster 3 |  |  |
| --- | --- | --- | --- | --- | --- | --- | --- | --- | --- | --- |
|  |  | F1 | Precision | AUC | F1 | Precision | AUC | F1 | Precision | AUC |
| Lockdown | Post-lockdown | 0.75 | 0.70 | 0.91 | 0.67 | 0.76 | 0.81 | 0.45 | 0.47 | 0.67 |
| Lockdown | Normal | 0.85 | 0.92 | 0.89 | 0.85 | 0.88 | 0.89 | 0.71 | 0.62 | 0.59 |
| Post-lockdown | Lockdown | 0.78 | 0.94 | 0.78 | 0.62 | 0.53 | 0.89 | 0.34 | 0.23 | 0.59 |
| Post-lockdown | Normal | 0.75 | 0.97 | 0.86 | 0.73 | 0.69 | 0.87 | 0.41 | 0.27 | 0.65 |
| Normal | Lockdown | 0.83 | 0.84 | 0.86 | 0.74 | 0.61 | 0.85 | 0.64 | 0.74 | 0.78 |
| Normal | Post-lockdown | 0.79 | 0.78 | 0.91 | 0.69 | 0.65 | 0.81 | 0.49 | 0.52 | 0.69 |

Given that the shrinkage discriminant analysis models for predicting cluster 3 performed poorly and that the PCA (Figure 2) showed a slight tendency for clusters 2 and 3 to merge, we decided to re-run the shrinkage discriminant analysis but remove cluster 3 as a predictive target. Importantly, this would indicate if the model metrics for predicting cluster 2 were negatively influenced by cluster 3 and the possible closer relationship between these two clusters relative to cluster 1. Repeating the shrinkage discriminant analysis indeed demonstrated that this was the case. Here, both the F1 and precision metrics significantly increased to about 0.90 for all iterations of training and testing datasets (Table 3).

**Table 3.** Performance metrics of the shrinkage discriminant analysis for binary classification of clusters 1 and 2 of mental health app users. Metrics reported include F1 and, in brackets, precision.

| Training Dataset | Testing Dataset | Cluster 1 vs. Cluster 2 |  |  |
| --- | --- | --- | --- | --- |
|  |  | F1 | Precision | AUC |
| Lockdown | Post-lockdown | 0.92 | 0.84 | 1 |
| Lockdown | Normal | 0.97 | 0.97 | 0.99 |
| Post-lockdown | Lockdown | 0.91 | 1.0 | 0.99 |
| Post-lockdown | Normal | 0.93 | 1.0 | 0.96 |
| Normal | Lockdown | 0.91 | 0.95 | 0.99 |
| Normal | Post-lockdown | 0.90 | 0.91 | 0.99 |

### Cluster 1 and cluster 2 of mental health app users represents two distinct groups of users who have either high or low mental well-being

Given that we found that clusters 1 and 2 were generalizable (i.e. could be predicted irrespective of the timepoint), we sought to identify the specific variables that characterize the app users in each cluster. To do this, we combined all three timepoint datasets into one, removed cluster 3, and used a Wilcoxon test with a Bonferroni correction to identify significant differences between the groups. Of the 46 included measures, 31 were significantly different between the two groups (Supplementary Table 1). Overall, cluster 1 was made up of users who self-identified as having high mental well-being. These users had low depression and suicidal ideation, a healthy quality of life, high well-being, and had a high level of perceived social support from family, friends, and significant others. They were also more likely to be female, have a paying job and high socioeconomic status, and more likely to identify their sexual orientation as heterosexual. Cluster 2, on the other hand, represented users who self-identified as having low mental well-being. They were more depressed and suicidal, had a poor quality of health, low well-being, and low levels of social support across all three domains. They were also more likely to be male, not have a paying job, low socioeconomic status, and more likely to identify as being LGBTQI+.

### Characterization of the unique features of cluster 3 of mental health app users

An interesting finding was that cluster 3 was not only poorly predicted but also that its removal from all three timepoints led to improved model performance for binary classification between clusters 1 and 2. As such, we then sought to identify the unique features that defined cluster 3 and how they may differ across timepoints. To do this, we first merged all three datasets (lockdown, post-lockdown, and normal) into a single dataset. We then narrowed down the features by determining the respective Pearson correlations between them and removing features with a r ≥ 0.75. Removing highly correlated features is an important step to reduce noise, therefore increasing computational power of the model, and increase the stability of the model. The correlation calculations indicated that six features met the criteria for exclusion: PSS1, PSS2, PSS4, PSS7, PSS9, and PSS10 (Supplementary Figure 1). All six features are questions from the Perceived Social Support scale and comprise part of the significant other (PSS1, PSS2, and PSS10), friends (PSS7 and PSS9), and family (PSS4) subscales. Importantly, the exclusion of these specific questions did not lead to the exclusion of the entire subscale for any of these measures.

After exclusion of the highly correlated features, 40 features remained. To estimate the contribution of features to the classification and regression trees (CART) model characterizing cluster 3, we performed recursive feature elimination (RFE). Here, the quality of the model was estimated on its ability to predict cluster 3 as a negative predictive value (NPV). Importantly, this method allows us to establish which, if any, features reliably predict cluster 3 membership irrespective of timepoint (i.e. are stable features of cluster 3). In doing so, we can also then identify those features that are not predictors, thereby establishing features of cluster 3 that may be unique to specific timepoints and warrant further investigation.

The RFE showed that using all 40 features, the CART model predicting cluster 3 had a high negative predictive value, suggesting that cluster 3 can indeed be predicted (Figure 3). Specifically, the RFE showed that 11 features are critical for the prediction of cluster 3. This is further confirmed by the fact that after 12 features, there is a clear plateau of NPV with a limited growth rate.

**Figure 3.**
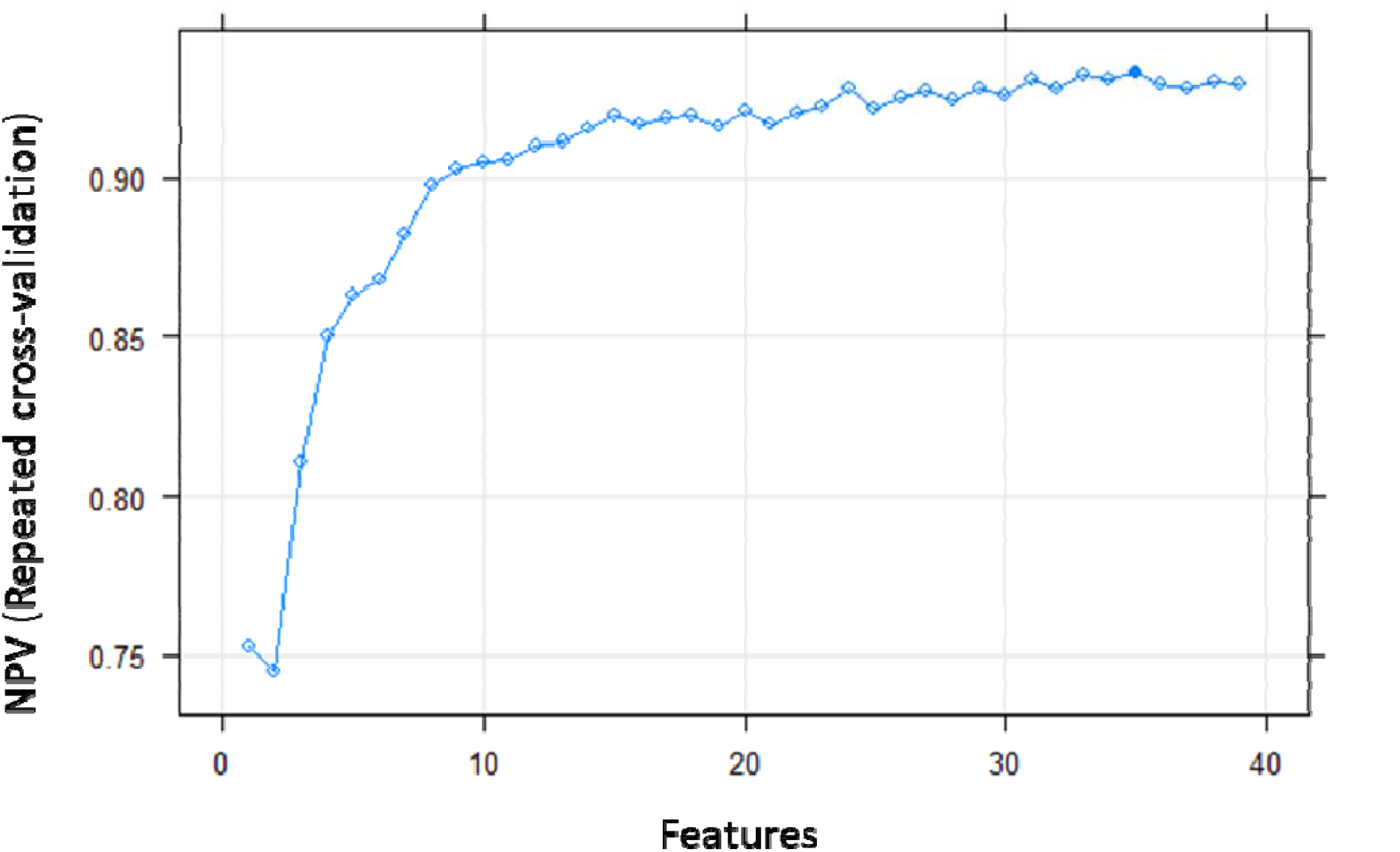
Plot showing the change in negative predictive value (NPV) based on the results of the recursive feature elimination (RFE). Here, we used RFE to estimate the contribution of features to the classification and regression trees (CART) characterizing cluster 3.

To further confirm our finding, we then performed two CARTs: one on the first 11 features and the second on the remaining 29 that fall after the initial plateau. Indeed, the first model with 11 features showed higher performance metrics relative to the second model (Table 4). Importantly, the NPV of the second model was 0.55, confirming our conclusion that the remaining 29 features are not important contributors to the overall prediction of cluster 3.

**Table 4.** Performance metrics of the classification and regression trees (CART) for two models: one comprising 11 features identified by the recursive feature elimination (RFE) as being important and one comprising the remaining 29 features that were identified by the RFE as being unimportant. PPV: positive predictive value; NPV: negative predictive value.

|  | Model 1 (11 features) | Model 2 (29 features) |
| --- | --- | --- |
| Metric |  |  |
| Sensitivity (recall) | 0.88 | 0.80 |
| Specificity | 0.68 | 0.48 |
| PPV (cluster 1&2) (Precision) | 0.86 | 0.75 |
| NPV (cluster 3) | 0.71 | **0.55** |
| F1 | 0.87 | 0.78 |

The 11 features that are critical for predicting cluster 3, irrespective of timepoint, were EQ-5D-5L6, KTEN, MED2, PSS3, PSS5, PSS6, PSS11, PSS12, WBS2, WBS3, and WBS7. We then used a Kruskall-Wallis followed by Dunn test to compare the 11 variables between cluster 3 and clusters 1 and 2 (Supplementary Table 2). Relative to the healthy cluster 1, cluster 3 had significantly higher depression, lower self-perception of health, mental health history, and poor well-being. They did, however, report similar levels of perceived social support to cluster 1, except that they rated family as less willing to help. Interestingly, despite cluster 3 having the same levels of depression and self-perception of health as the unhealthy cluster 2, they reported higher levels of social support. Cluster 3 users, however, were more likely to have a history of mental health concerns than those in cluster 2.

We then investigated the nature of the 29 remaining features for cluster 3 across the three individual timepoints, again using a Kruskall-Wallis test followed by a Dunn test for pairwise comparisons (Table 5). Strikingly, this showed that cluster 3 was almost identical to the unhealthy cluster 2 during lockdown, with only one variable, PSS8, reaching a very small statistically significant difference (p = 0.048) between the two groups. In the normal timepoint, however, cluster 3 was identical to healthy cluster 1 on all 29 variables. Statistical analysis of cluster 3 during post-lockdown revealed a unique group that fit somewhere in between cluster 2 and 3. Relative to cluster 2, cluster 3 users were more likely to be male, drink alcohol, smoke tobacco, and talk about problems with their family. Relative to cluster 1, cluster 3 were more likely to have previous contact with hospitals and clinics for mental health services, take prescribed medicine for mental health, and smoke tobacco. They were also less likely to have a paying job, engage with online mental self-help resources, and feel that they were dealing with problems and thinking clearly.

**Table 5.**
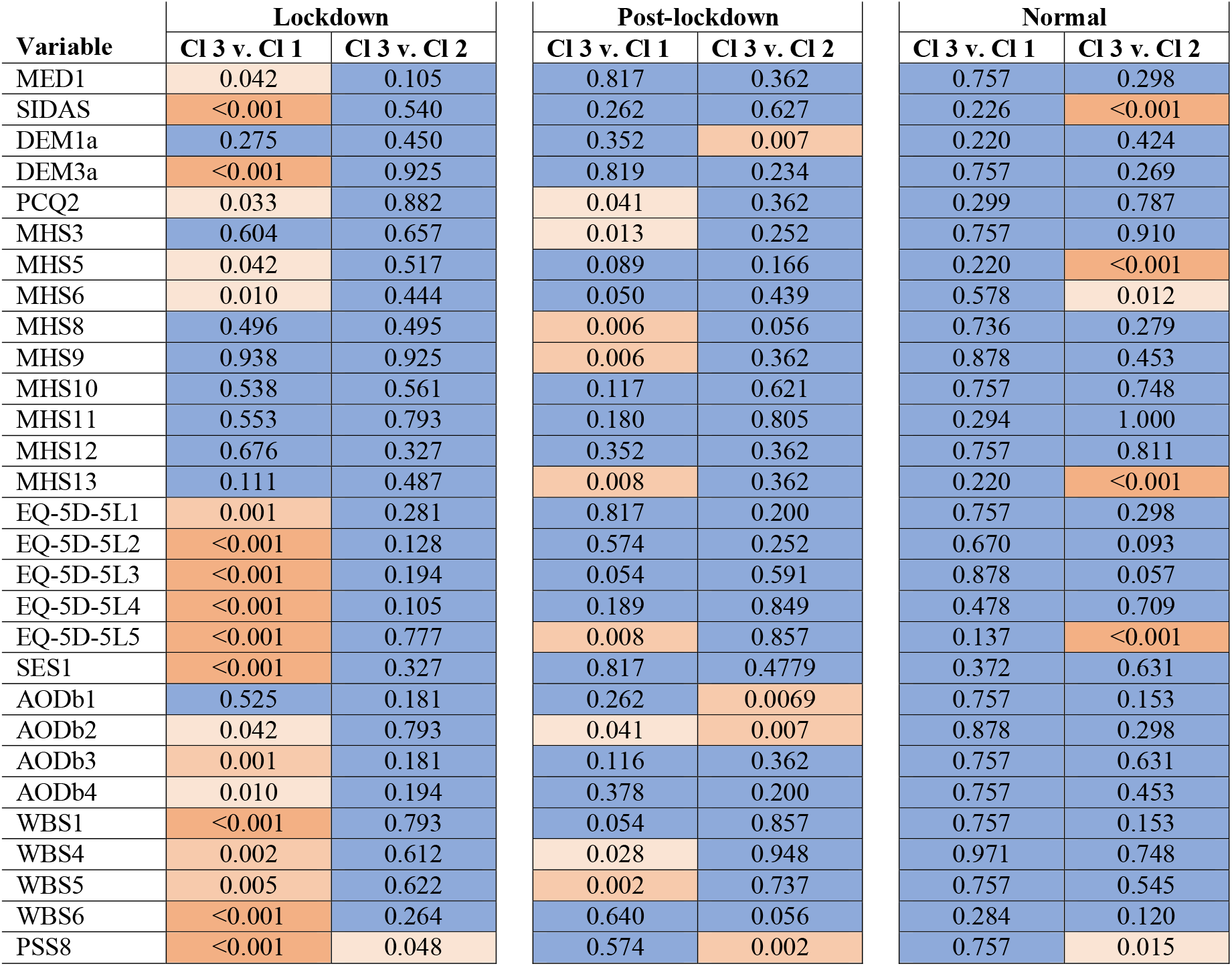
Pairwise comparisons of the 29 features for cluster 3 across the three individual timepoints. Orange color indicates statistically significant and blue indicates no statistically significant effect.

### Characterization of cluster 3 across COVID-19 timepoints

The characterization of cluster 3 mental health app users indicated that there were some stable features of cluster 3. Most of their features, however, alternated between being similar to the other clusters in lockdown and normal timepoints, respectively, and to having very unique features post-lockdown. As such, we suspected that this may underlie our inability to predict cluster 3 in our initial machine learning model. To confirm this, we performed a PCA of the three timepoints to determine how they compared to one another based on user characteristics (Figure 4A). The PCA demonstrated that users’ characteristics in each of the three clusters were more similar to one another in the lockdown and normal timepoints and that the post-lockdown timepoint was different for all users. We further confirmed the PCA finding with a cluster dendrogram analysis that also showed that lockdown and normal timepoints are more similar to one another than post-lockdown (Figure 4B; Supplementary Table 3).

**Figure 4.**
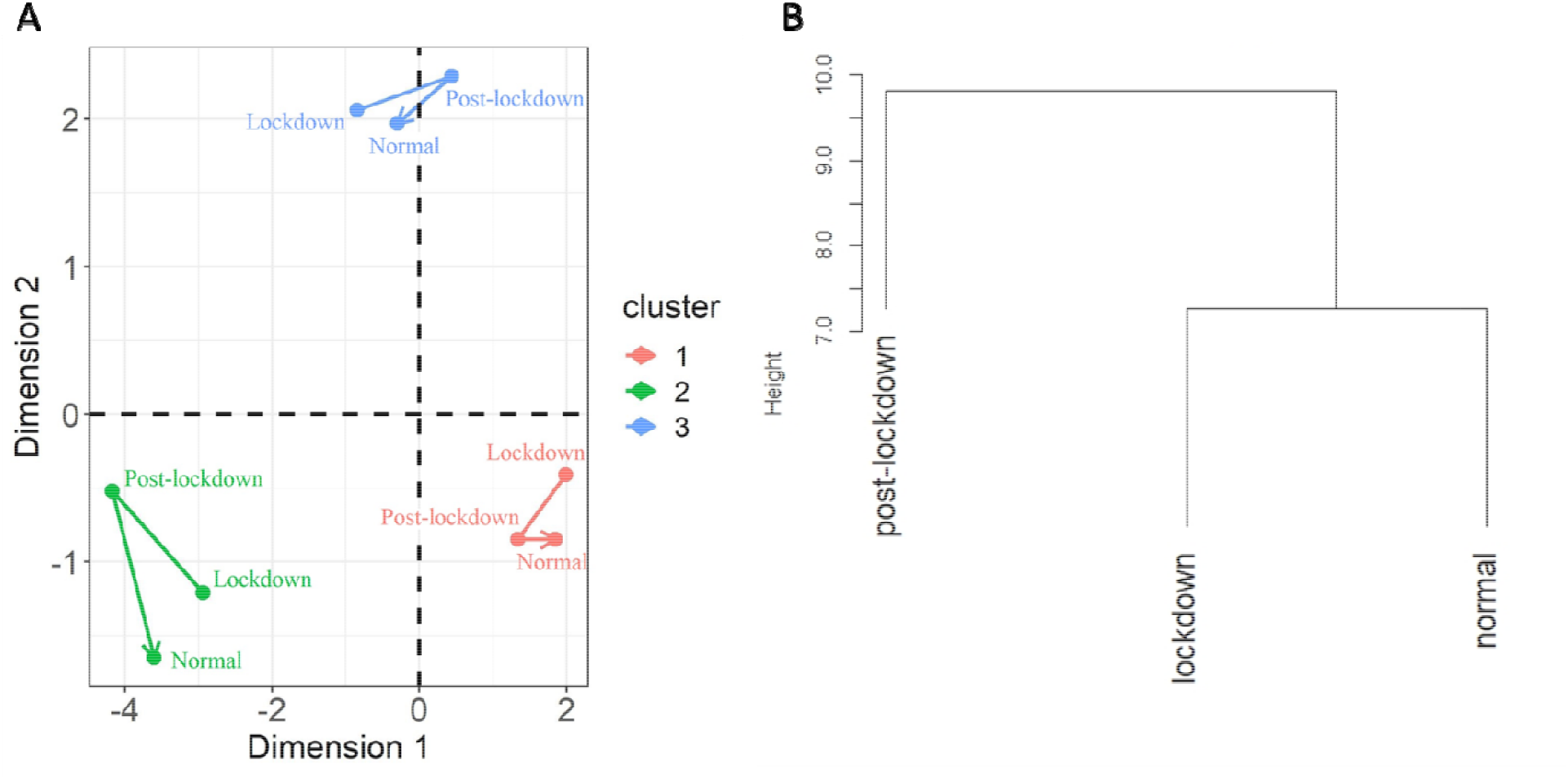
Analysis of cluster 3 characteristics across the three COVID-19 timepoints. (A) Principal component analysis (PCA) dot plot. (B) Cluster dendrogram.

## Discussion

Using the screening data from a novel mental health app, we were able to identify that there were three distinct subgroups of users that differed in their demographic and mental health characteristics. A combination of k-means clustering and PCA indicated that all COVID-19 timepoints were consistently characterized by two of the three subgroups: *lower mental well-being* and *higher mental well-being*. These two distinct subgroups of app users were consistent across all the COVID-19 timepoints, suggesting that significant external life stressors (e.g. lockdown) had a limited effect on changing the fundamental characteristics that defined these subgroups. The lower mental well-being subgroup was characterized by users with higher depression and suicidality, poorer quality of health, and lower levels of social support. Users in this subgroup were more likely to be male and identify as being LGBTQIA+, not have a paying job, and have a lower socioeconomic status. These characteristics are well-documented as known factors to be associated with poor mental health outcomes in young people ^31, 32^.

An interesting, and somewhat unexpected, finding of the current study was the subgroup of users who had consistently higher mental well-being. The higher mental well-being subgroup was made up of users with lower depression and suicidal ideation, healthier quality of life, and higher level of perceived social support. These users were also more likely to identify as heterosexual females, have a paying job and high socioeconomic status. To our knowledge, there is no literature examining the use of health apps among people who may already have higher levels of mental well-being or health. Further, there is some contention about whether healthier people should even use health intervention apps. While healthier people may benefit from encouragement and advice about remaining healthy, they may become more anxious about their health from using an intervention app, resulting in a paradoxical effect of the app decreasing health and well-being ^33^. Our finding highlights the possibility that there may be a subgroup of mental health app users who are seeking to maintain their current mental well-being, rather than having ongoing issues that they are seeking to remedy. Further, these users may have different goals and requirements to someone with current mental health concerns, suggesting that JITAI apps and personalized interventions need to address this.

Future research, therefore, would benefit from including questions that assess whether a user may be interested in simply maintaining their mental well-being. It is worth noting, however, that the higher mental well-being subgroup may still have poorer mental health and well-being relative to someone who does not access a mental health app. This highlights the importance of including a group of people who choose not to access mental health apps in future research. In doing so, we will be able to assess the status of this higher mental well-being subgroup relative to both the lower mental well-being subgroup and those who do not need, or chose to use, these apps.

Although there was a third subgroup of users identified in all three timepoints, our supervised machine learning was unable to predict them, highlighting that the characteristics of this group were highly influenced by COVID-19. Our CART and RFE identified that only 11 characteristics were stable across time in this subgroup: low self-rated health, high depression, higher likelihood of having mental health history, and low mental well-being. Interestingly, this subgroup reported consistently high levels of perceived social support except that they rated family as less willing to help. The remaining characteristics of this subgroup were variable across the COVID-19 timepoints. During the COVID-19 lockdown, these users were identical to the lower mental well-being subgroup. Three months after the COVID-19 lockdown (normal timepoint), this subgroup’s remaining characteristics became identical to the higher mental well-being group. This third subgroup, however, fit somewhere in between the higher and lower mental well-being subgroups during the period immediately following the COVID-19 lockdown (post-lockdown timepoint). Relative to the lower well-being subgroup, these users were more likely to be male, drink alcohol, smoke tobacco, and talk about problems with their family. Relative to the higher mental well-being subgroup, these users were more likely to report previous contact with hospitals and clinics for mental health services, take prescribed medicine for mental health, and smoke tobacco. They were also less likely to have a paying job, engage with online mental self-help resources, and feel that they were dealing with problems and thinking clearly. These findings suggest that this subgroup’s mental well-being is determined by the situation, and we therefore entitled this group the *situational mental well-being* subgroup.

Based on our results, it appears that this situational mental well-being subgroup specifically is highly influenced by traumatic stressors and stressful situations. In situations of high stress (COVID-19 lockdown) this subgroup mirrors the lower mental well-being subgroup. In situations of low stress (normal timepoint), this subgroup bears greater resemblance to the higher mental well-being subgroup, although they do still report symptoms of depression, low mental well-being, and poor health, albeit to a lesser extent than the lower subgroup. In the period immediately after the traumatic stressor (post-lockdown), the situational mental well-being subgroup has high depression and low mental well-being. They appear to engage in more maladaptive coping behaviours, including increased drinking and smoking, as well as report increased use of psychiatric medication. They also report that they feel they are not dealing with problems and thinking clearly and report that they are talking about their problems with family. This set of behaviours and feelings are associated with burnout in university students.

Burnout is characterized by the presence of exhaustion, cynicism, and inefficacy, and is associated with symptoms including depression and low mental well-being ^34, 35^. Studies have reported that burnout in university students is highly associated with smoking, problem drinking, and substance use/abuse ^36, 37, 38^. Further, another study found that in addition to these behaviours, university students were also more likely to seek out support from their family ^36^. To date, there has been little research into the effects of different COVID-19 stages on mental health and burnout in university students and no research on how this affects mental health app users. Generally, studies have pointed to increased burnout in university students across the COVID-19 pandemic ^8, 39, 40^. One study suggested that burnout rates occurred in those students with higher levels of existing depressive symptoms, a finding that supports our own ^39^. Interestingly, another study found that burnout progressively increased across a one-year period of COVID-19 ^8^. This may account for our finding that post-lockdown was associated with symptoms indicative of burnout and suggests that the effects (burnout) of a prolonged traumatic stressor persist after the stressor is removed. The presence of the situational mental well-being subgroup that appears to be susceptible to burnout highlights that future studies on mental health apps would benefit from including a measure for burnout, such as the Maslach Burnout Inventory for Students (MBI-SS) ^41^. It may also be the case that those accessing mental health apps for burnout symptoms have different needs and is therefore an important consideration for JITAI-style personalized interventions.

Despite the strengths of the paper, there are some limitations. First, there were differences in the number of users who completed screening for the app across the three COVID-19 timepoints. Specifically, there were more users during the COVID-19 lockdown and therefore more datapoints in the lockdown dataset relative to the other two timepoints. Studies have reported that during COVID-19 lockdowns, mental health app usage increased as people sought out pandemic-related stress coping techniques ^42^. Despite this, we still found that irrespective of COVID-19 timepoints, two subgroups had stable user characteristics, which were further validated with machine learning and alternating training and testing datasets. This suggests that the imbalance in the number of users between the datasets had little impact on the study results and conclusions. It is also important to note that the users of our mental health app do not report formal clinical diagnosis for their mental health concerns (e.g. clinician diagnosed depression). Future research, therefore, would also benefit from examining how a formal mental health diagnosis may affect user characteristics. There is, however, still an important need for people to access mental health irrespective of formal diagnoses. For example, university students may experience a brief period where they have poor mental well-being and high distress and need a brief self-help intervention to help them address this. Our study highlights that even in this self-help-directed user group, there are still unique characteristics that may need to be catered for in JITAI mental health apps.

## Conclusion

Smartphone-based mental health interventions have significantly grown in popularity and have been shown to improve user mental health, especially among university students. Despite this, overall user engagement has remained low. In an effort to combat this, recent studies have shown that users can be subgrouped based on specific characteristics (e.g. gender). To date, however, no studies have examined how the extreme environmental stressors of the COVID-19 pandemic and its associated lockdowns have influenced these subgroups. Using machine learning, we demonstrated that there are three unique subgroups of university students who access mental health apps. Two of these, the higher well-being and lower well-being groups, were consistent over the COVID-19-related timepoints, suggesting that they are both stable groups of users irrespective of external environmental factors. The characteristics of the third subgroup (situational well-being), however, were highly dependent on the COVID-19 timepoint and showed feelings and behaviours consistent with burnout. This suggests that the situational well-being subgroup is highly influenced by traumatic stressors and stressful situations. Overall, these findings clearly demonstrate that users have different characteristics and therefore likely have different goals from mental health app use. Although this highlights the importance of JITAI-style personalized intervention apps, it may also be important for these apps to include explicit questions asking the user why they are using the app when allocating them to a particular intervention. Further, these apps may also benefit from including assessments of, and interventions for, burnout and its associated symptoms. Overall, these approaches are likely to improve mental health app user engagement among university students.

## Data Availability

Data may be made available on request and subject to the relevant governance procedures.

## Funding Statement

This work was supported by Commonwealth of Australia Medical Research Future Fund grant MRFAI000028 Optimising treatments in mental health using AI

## Conflicts of Interest

The authors declare no conflicts of interest.

## Data Availability

Data may be made available on request and subject to the relevant governance procedures.

## Author Contributions

A.S. Conceived and designed the study, performed the machine learning and data analyses, and wrote the manuscript.

L.H., K.M., S.G., and S.V. assisted with data collection and co-wrote the manuscript.

A.W., S.K., W-Y.Z., J.B., O.M., J.H., H.C., and J.N. assisted with conceptualization and co-wrote the manuscript.

## Notes

### Competing Interest Statement

The authors have declared no competing interest.

### Author Declarations

The study was approved by the University of New South Wales Human Research Ethics Committee, approval no. HC200466

