## Supplementary Methods and Figure for "Machine learning identifies a COVID-19-specific phenotype in university students using a mental health app"

**Supplementary Material**

**Supplementary Methods: Psychological Questionnaires**

*Abridged NIDA-Modified ASSIST Drug Screening Tool (AOD)*: This scale measures substance use. In the abridged version, responders are asked about the number of alcoholic drinks consumed a day, consumption of tobacco products, and non-medical prescription and illegal drug use. Responses are scored on a 5-point Likert scale ranging from 1 (never used) to 5 (daily use) ^1^.

*Demographics*: A series of demographic questions were asked including self-identified gender, sex at birth, and sexual orientation. For gender and sex at birth, response options were: female, male, non-binary, use of a different term, and prefer not to say. Sexual orientation, response options were: heterosexual, gay or lesbian, bisexual, use of a different term, don’t know, prefer not to say.

*EuroQuol-5D 5-level version (EQ-5D-5L)*: A five-item measurement of health including: mobility, self-care, usual activities, pain/discomfort, and anxiety/depression. Each item is rated on a 3-point scale: no problems, some problems, and extreme problems ^2^.

*Kessler Psychological Distress Scale, 10-item version (KTEN)*: This scale assesses non-specific psychological distress, including depression, anxiety, and worry. Questions are scored using a 5-point scale ranging from 1 (none of the time) to 5 (all of the time) ^3^.

*Use of Mental Health Care Services (MHS)*: The MHS assesses a person’s use of professional mental health services in the past 12 weeks, such as hospital services, allied health services, and community-based services. Level of use is rated on a 7-point scale ranging from 0 (0 times) to 6 (more than 5 times) ^4^.

*Perceived Stress Scale (PSS)*: The PSS measures the perception of life stress, with subscales assessing experiences of stress that feels unpredictable, uncontrollable, and overwhelming. Items are rated using a 5-point Likert scale ranging from 0 (never) to 4 (very often) ^5^.

*Physical and Mental Health (MED)*: This measure consisted of two questions regarding whether participants had a previous diagnosis for either a chronic physical health condition or a mental health condition. Responses were: no, yes, unsure, or prefer not to say.

*Productivity Cost Questionnaire (PCQ):* The PCQ measures health-related productivity losses using a range of scales including open choice, 11 item Likert, numeric fill in, two item closed choice, and 5-item multiple choice ^6^.

*Suicidal Ideation Attributes Scale (SIDAS)*: This measure assesses the level of suicidal thoughts and ideation. The final total score of this scale ranges from 0 (low) to 50 (extremely high) ^7^.

*Short Warwick Edinburgh Mental Well-being Scale (WBS)*: The WBS is a short scale that measures mental well-being. Each item on this scale is scored using a 5-point Likert scale ranging from 1 (none of the time) to 5 (all of the time) ^8^.

*Subjective Socioeconomic Status Scale (SES)*: This measure assesses socioeconomic status. It uses a ladder metaphor to represent the relative success (more or less) that an individual perceives that they have ^9^.

1.	National Institute on Drug Abuse. NIDA Drug Screening Tool, NIDA-Modified ASSIST (NM ASSIST).). National Institutes of Health (2020).

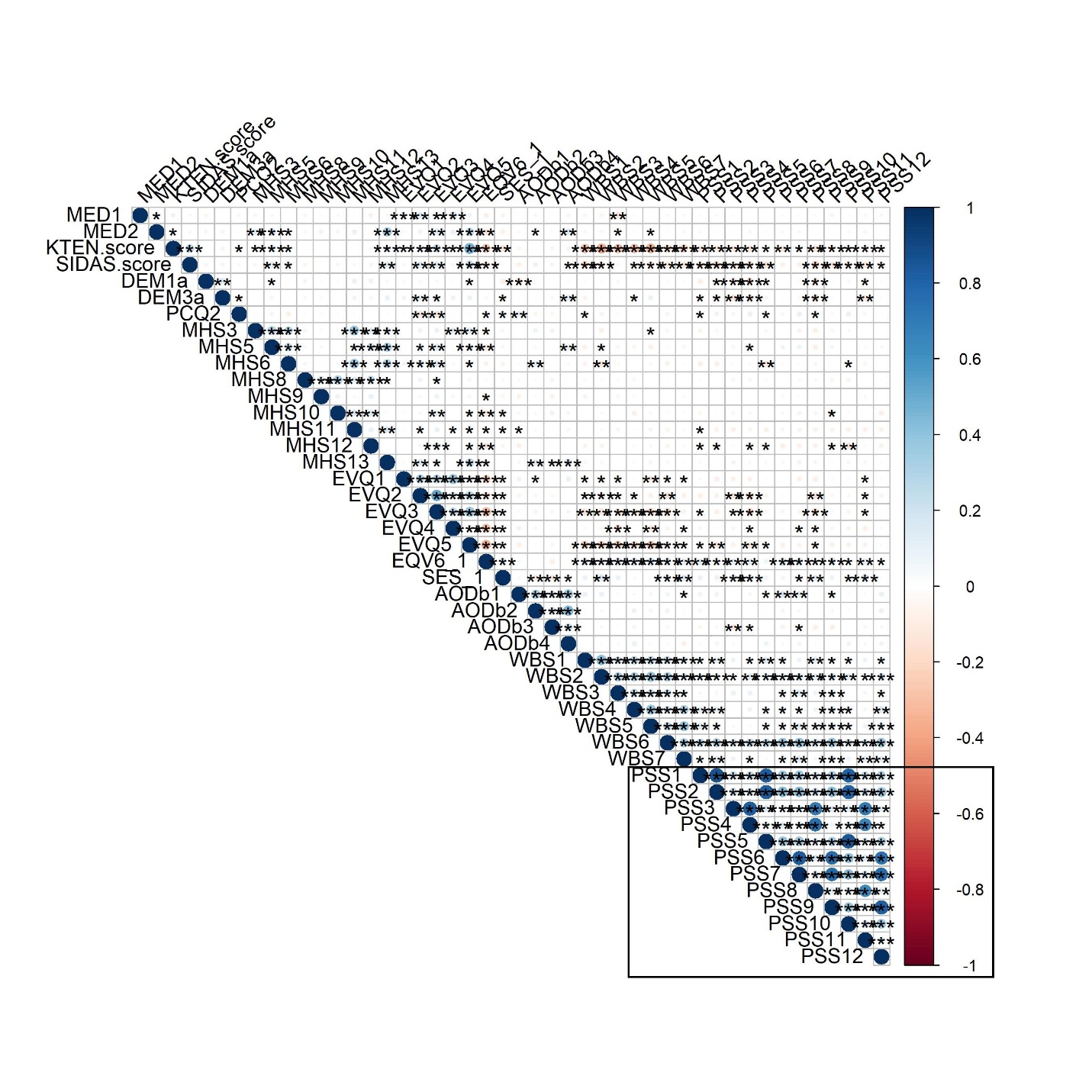

**Supplementary Figure 1.** Pearson correlations between features within a single merged dataset
