## Supplementary Tables for "Machine learning identifies a COVID-19-specific phenotype in university students using a mental health app"

**Supplementary Table 1.** Comparison of specific variables that characterize clusters 1 and 2

| Var | statistic | p-value | general.adj.p.value |
| --- | --- | --- | --- |
| MED1 | 6271.5 | 0.44259 | 0.522 |
| MED2 | 5979 | 0.887973 | 0.888 |
| KTEN score | 2976.5 | 2.13E-10 | 0 |
| SIDAS score | 3896 | 1.93E-07 | 0 |
| DEM1a | 5222 | 0.00598 | 0.011 |
| DEM3a | 4489 | 0.000229 | 5.00E-04 |
| PCQ2 | 6936.5 | 0.006411 | 0.0113 |
| MHS3 | 5795.5 | 0.171222 | 0.2188 |
| MHS5 | 5607 | 0.304801 | 0.3789 |
| MHS6 | 5687 | 0.13473 | 0.1823 |
| MHS8 | 5884 | 0.699614 | 0.7662 |
| MHS9 | 5960 | 0.85338 | 0.8765 |
| MHS10 | 5978 | 0.772419 | 0.8263 |
| MHS11 | 6012 | 0.85743 | 0.8765 |
| MHS12 | 5352 | 0.030854 | 0.0473 |
| MHS13 | 5858 | 0.64961 | 0.7288 |
| EQV1 | 5672.5 | 0.039547 | 0.0587 |
| EQV2 | 5225 | 0.011037 | 0.0181 |
| EQV3 | 4693 | 0.001886 | 0.0038 |
| EQV4 | 5669.5 | 0.36759 | 0.445 |
| EQV5 | 3872.5 | 1.23E-06 | 0 |
| EQV6 | 8727 | 2.14E-08 | 0 |
| SES1 | 7419 | 0.003672 | 0.007 |
| AODb1 | 6799.5 | 0.098366 | 0.1371 |
| AODb2 | 5426 | 0.066085 | 0.095 |
| AODb3 | 5754.5 | 0.139681 | 0.1836 |
| AODb4 | 5816 | 0.525352 | 0.6042 |
| WBS1 | 7934.5 | 1.04E-05 | 0 |
| WBS2 | 8290.5 | 1.26E-07 | 0 |
| WBS3 | 7058 | 0.016839 | 0.0267 |
| WBS4 | 7864 | 1.54E-05 | 0 |
| WBS5 | 7742 | 5.65E-05 | 1.00E-04 |
| WBS6 | 8675 | 5.77E-09 | 0 |
| WBS7 | 7183 | 0.010685 | 0.0181 |
| PSS1 | 11277 | 2.69E-28 | 0 |
| PSS2 | 11538.5 | 2.93E-31 | 0 |
| PSS3 | 8974 | 3.50E-10 | 0 |
| PSS4 | 9080 | 8.85E-11 | 0 |
| PSS5 | 11086.5 | 2.23E-26 | 0 |
| PSS6 | 9573 | 6.12E-14 | 0 |
| PSS7 | 9680 | 1.26E-14 | 0 |
| PSS8 | 9182 | 3.02E-11 | 0 |
| PSS9 | 9818.5 | 6.39E-16 | 0 |
| PSS10 | 11410 | 6.36E-30 | 0 |
| PSS11 | 9177 | 1.16E-11 | 0 |
| PSS12 | 9543.5 | 9.27E-14 | 0 |

**Supplementary Table 1.** Comparison of 11 variables between clusters 3 and clusters 1 and 2

| Var | statistic | p-value | first - second - p-value | first - third_pl1 - p-value | second - third_pl1 - p-value | general.  adj.p.value | first_second_  adj.p.value | first_third_  adj.p.value | second_third_  adj.p.value |
| --- | --- | --- | --- | --- | --- | --- | --- | --- | --- |
| MED2 | 15.22535 | 0.000494 | 0.90923 | 0.000212 | 0.001631 | 0.000494 | 0.90923 | 0.000292 | 0.003493 |
| KTEN | 64.80258 | 8.48E-15 | 1.43E-10 | 1.33E-11 | 0.704174 | 1.97E-14 | 3.93E-10 | 3.65E-11 | 0.704174 |
| EQV6 | 58.21042 | 2.29E-13 | 2.79E-08 | 9.44E-12 | 0.24346 | 3.60E-13 | 4.38E-08 | 3.46E-11 | 0.297563 |
| WBS2 | 47.00157 | 6.22E-11 | 1.73E-07 | 2.57E-09 | 0.485412 | 8.55E-11 | 2.37E-07 | 5.66E-09 | 0.533953 |
| WBS3 | 21.90493 | 1.75E-05 | 0.01661 | 4.27E-06 | 0.050221 | 1.93E-05 | 0.018271 | 6.72E-06 | 0.069053 |
| WBS7 | 24.34054 | 5.18E-06 | 0.010774 | 1.29E-06 | 0.0416 | 6.33E-06 | 0.013168 | 2.37E-06 | 0.065372 |
| PSS3 | 101.1186 | 1.10E-22 | 1.72E-08 | 1.80E-22 | 0.000241 | 4.04E-22 | 3.16E-08 | 1.83E-21 | 0.000663 |
| PSS5 | 134.9154 | 5.05E-30 | 1.38E-26 | 0.50949 | 9.02E-23 | 5.56E-29 | 1.52E-25 | 0.560439 | 9.92E-22 |
| PSS6 | 64.6884 | 8.98E-15 | 1.68E-14 | 0.719282 | 2.43E-10 | 1.97E-14 | 9.22E-14 | 0.719282 | 1.34E-09 |
| PSS11 | 103.52 | 3.32E-23 | 4.71E-10 | 3.32E-22 | 0.001905 | 1.83E-22 | 1.04E-09 | 1.83E-21 | 0.003493 |
| PSS12 | 59.846 | 1.01E-13 | 3.77E-14 | 0.277845 | 2.09E-08 | 1.85E-13 | 1.38E-13 | 0.339588 | 7.67E-08 |

**Supplementary Table 3.** Euclidean distance between clusters at specific timepoints

| Cluster | 1 | 2 | 3 | 1 | 2 | 3 |
| --- | --- | --- | --- | --- | --- | --- |
|  | Post-lockdown | | | Normal | | |
| Lockdown | 0.78 | 1.41 | 1.31 | 0.46 | 0.79 | 0.56 |
| Post-lockdown | N/A | N/A | N/A | 0.51 | 1.27 | 0.81 |
